# Muscle pathology of antisynthetase syndrome according to antibody subtypes

**DOI:** 10.1101/2022.04.25.22274260

**Authors:** Jantima Tanboon, Michio Inoue, Shinya Hirakawa, Hisateru Tachimori, Shinichiro Hayashi, Satoru Noguchi, Naoko Okiyama, Manabu Fujimoto, Shigeaki Suzuki, Ichizo Nishino

**Affiliations:** Department of Neuromuscular Research, National Institute of Neuroscience, National Center of Neurology and Psychiatry (NCNP), Tokyo 187-8502, Japan; Department of Genome Medicine Development, Medical Genome Center, National Center of Neurology and Psychiatry (NCNP), Tokyo 187-8502, Japan; Department of Clinical Data Science, Clinical Research & Education Promotion Division, National Center of Neurology and Psychiatry (NCNP), Tokyo 187-8502, Japan; Department of Dermatology, Graduate School of Medical and Dental Sciences, Tokyo Medical and Dental University, Tokyo 113-8510, Japan; Department of Dermatology, Faculty of Medicine, University of Tsukuba, Ibaraki 305-8576, Japan; Department of Dermatology, Graduate School of Medicine, Osaka University, Osaka 565-0871, Japan; Department of Neurology, Keio University School of Medicine, Tokyo 160-8582, Japan; Department of Clinical Genome analysis, Medical Genome Center, National Center of Neurology and Psychiatry (NCNP), Tokyo, 187-8502 Japan

**Keywords:** antisynthetase, anti-tRNA synthetase, myositis, muscle pathology, HLA-DR

## Abstract

Antisynthetase syndrome is recently recognized as one of the major entities of autoimmune myositis. The prototype of antisynthetase syndrome is anti-Jo-1 antibody associated syndrome while the syndromes associated with non-Jo-1 antisynthetase antibodies are clinically and pathologically less recognized. Identifying a non-Jo-1 antisynthetase syndrome patient could be challenging because the full panel serology test may not be available at the time of diagnosis in addition to technical difficulty especially for anti-OJ antibody detection. This study aimed to characterize the muscle pathology and explore the utility of myofiber HLA-DR expression for the diagnosis of antisynthetase syndrome.

We retrospectively compared 212 muscle biopsies from antisynthetase syndrome patients regarding four pathology domains and histology of interests using *t* test and Fisher’s exact test as appropriate. We further compared the myofiber HLA-DR expression pattern in antisynthetase syndrome with 602 muscle biopsies with other autoimmune myositis and 140 muscle biopsies with other myopathies potentially containing myositis-like pathology and calculated sensitivity, specificity, positive predictive value, and negative predictive value to identify the most diagnostic pattern for antisynthetase syndrome.

The most common myopathological pattern in antisynthetase syndrome was necrotizing myopathy (46.2%). Perifascular necrosis was present in 28.3% of antisynthetase syndrome. Anti-OJ and anti-EJ antisynthetase syndrome were associated with high muscle fiber scores. Anti-OJ also showed high inflammatory domain score. If muscle biopsies suspicious for dermatomyositis by sarcoplasmic myxovirus resistance protein A immunohistochemical expression and those with inclusion body myositis clinicopathology were excluded, myofiber HLA-DR expression was the most diagnostic of antisynthetase syndrome with 95.4% specificity, 61.2% sensitivity, 85.9% positive predictive value, and 84.2% negative predictive value. HLA-DR expression in perifascicular fibers was highly specific to anti-Jo-1 antisynthetase syndrome.

Anti-OJ antisynthetase syndrome has more prominent myopathology than the other antisynthetase syndrome subtypes. Presence of myofiber HLA-DR expression in a clinicopathologically approved non-dermatomyositis and non-inclusion body myositis muscle biopsy is highly indicative of antisynthetase syndrome. Presence of HLA-DR expression suggests the involvement of type II interferon in the pathogenesis in antisynthetase syndrome subpopulation although the detailed mechanism and the reason for preferential perifascicular localization are yet to be identified.

## Introduction

Antisynthetase syndrome (ASS) is an entity diagnosed serologically by the presence of one of eight clinically associated anti-aminoacyl transfer RNA synthetase (antisynthetase, anti-ARS) autoantibodies, namely, anti-Jo-1 (histidyl), anti-PL-7 (threonyl), anti-PL-12 (alanyl), anti-EJ (glycyl), anti-OJ (isoleucyl), anti-KS (asparaginyl), anti-Ha (tyrosyl), and anti-Zo (phenylalanyl), in addition to various combinations of clinical features, including myositis, interstitial lung disease (ILD), skin lesions such as mechanic’s hands, arthritis/arthralgia, fever, and Raynaud phenomenon.^1^ Anti-Jo-1-associated ASS is the first clinically and pathologically characterized ASS subtype.^1-3^ Anti-Jo-1 is the most common anti-ARS antibody as well as the most common myositis-specific antibody in autoimmune myositis (also called idiopathic inflammatory myopathy). The anti-Jo-1 antibody was present in 18.7% of the patients in a large cohort of 1637 patients with autoimmune myositis from the EuroMyositis registry, while the other six clinically associated non-Jo-1 ARS antibodies (excluding anti-Ha) were collectively found in 3.5% of the cases (0.2%-1.3% for each antibody subtype).^4^ Notably, among 828 patients in a collaborative study cohort of ASS from the American and European Network of Antisynthetase Syndrome (AENEAS) collaborative group, the prevalence of anti-Jo-1 ASS was 72%, while the prevalence of each non-Jo-1 ASS (including anti-OJ, anti-PL-7, anti-PL-12, and anti-EJ ASS) ranged from 2% to 11.5%.^5^ The small number of studies focusing on the clinical features of non-Jo-1 ASS is likely due to their much lower prevalence. Thus, studies on the myopathological features of non-Jo-1 ASS are rare.

Historically, without serological information, the application of Bohan and Peter^6^ classification is likely to categorize ASS as polymyositis (PM) and, in the patients with skin lesions, as dermatomyositis (DM). Notably, an “ASS corresponding cluster” categorized by a hierarchical cluster analysis based on clinical manifestations and myositis-specific antibodies was composed of 95% and 5% of cases previously diagnosed as PM and DM, respectively).^7^ The application of subsequently proposed classification systems without serological information also risks misclassification of ASS as DM and vice versa because of overlapping clinicopathological features.^8^ The presence of features suggestive of DM diagnosis, including DM skin lesions (i.e., Gottron signs/papules and/or heliotrope rashes) and perifascicular atrophy (PFA),^9^ has been reported in 15%-28%^10-11^ and 44.4%^12^ of the patients at the time of ASS diagnosis, respectively. In contrast, the features proposed as ASS diagnostic criteria, including mechanic’s hands and ILD,^13-14^ are commonly present in anti-MDA5 DM and have been reported in 42.7% and 76.5% of a cohort of 121 anti-MDA5 DM, respectively.^15^ Since complete serological results may not be available at the time of muscle pathology evaluation, we aimed to identify the characteristic myopathological features of ASS to help identify patients in such situations. We chose to explore myofiber HLA-DR expression as a possible diagnostic feature for ASS because (1) other histochemical features previously described in ASS can be present in non-ASS patients, especially those with DM.^16^ ; (2) HLA-DR has been reported mainly in inclusion body myositis (IBM) and ASS and less commonly in other autoimmune myositis and non-autoimmune myositis entities.^12,17,18^ ; and (3) peculiar perifascicular patterns have been reported in anti-Jo-1 ASS.^12, 19^

## Materials and Methods

### Patients

This study included muscle biopsies from 212 myositis patients who were serologically positive for any of the anti-ARS antibodies and whose muscle biopsies were sent to the National Center of Neurology and Psychiatry (NCNP), a nation-wide referral center for muscle disease, for pathological evaluation for the purpose of diagnosis between January 2009 and September 2019 (n = 212: anti-Jo-1 = 65, anti-OJ = 20, anti-PL-7 = 20, anti-PL-12 = 11, anti-EJ = 10, anti-KS = 1, and anti-ARS positive, not otherwise specified = 85). This study was an expansion of the ASS cohort in our previous studies.^20-22^ For comparison, we included 602 muscle biopsies from patients with other well-established subtypes of autoimmune myositis, i.e., dermatomyositis (DM, n = 188: anti-TIF1-γ = 65, anti-Mi-2 = 30, anti-MDA5 = 22, anti-NXP-2 = 60, anti-SAE = 5, and seronegative DM = 6), immune-mediated necrotizing myopathy (IMNM, n = 313: anti-SRP = 188 and anti-HMGCR = 125), in addition to 140 muscle biopsies from patients with genetically confirmed myopathies that are known to show inflammatory features to some extent^23, 24^ categorized as “possible myositis mimics” (P-MM, including dysferlinopathy = 50, sarcoglycanopathy = 15, laminopathy = 16, anoctamin-5 (ANO5) myopathy = 3, fukutin-related protein (FKRP) myopathy = 9, and facioscapulohumeral muscular dystrophy (FSHD) = 47) and recently diagnosed inclusion body myositis (IBM, between January 2019 and September 2019, n = 101). Patients were classified as adult if they were ≥ 18 years old.

### Serological information and inclusion criteria for autoimmune myositis

Anti-ARS and DM-specific antibodies were identified using previously described methods.^16^ ASS was defined as seropositivity for one of the eight anti-ARS antibodies and classified according to the antibody subtype. Patients who tested positive on an ELISA using recombinant ARS antigens (Jo-1, PL-7, PL-12, EJ, and KS) but underwent no further tests to specify the antibody subtype were categorized in the “anti-ARS positive, not otherwise specified” subtype (anti-ARS_NOS). All anti-OJ ASS cases were identified by RNA immunoprecipitation; seven cases were selectively tested among patients who tested negative for other myositis-specific antibodies but showed the clinicopathological impressions of autoimmune myositis.

The diagnostic criteria for DM in this study was sarcoplasmic expression for myxovirus resistance protein A (MxA) immunohistochemistry and positivity for one of five DM-specific antibodies (anti-TIF1-γ, anti-Mi-2, anti-MDA5, anti-NXP-2, and anti-SAE) or negative results for all DM-specific antibodies; the latter was classified as seronegative DM.^16^

For IMNM, we included patients with positive results for either anti-SRP or anti-HMGCR autoantibodies. In Japan, anti-SRP antibodies were detected by RNA immunoprecipitation and/or ELISA, while anti-HMGCR antibodies were detected by protein immunoprecipitation and/or ELISA as described previously.^25,26^ Anti-SRP autoantibodies were also recognized by commercialized immunoblot.

We used the Llyod et al^27^ criteria to identify IBM and also used the patchy/dot-like pattern of p62 as a surrogate marker for rimmed vacuoles.

### Possible myositis mimics (P-MM)

Genetic diagnoses of dysferlinopathy, sarcoglycanopathy, laminopathy, ANO5 myopathy, and FKRP myopathy were made using targeted resequencing gene panels, as described previously.^28^ Notably, all dysferlinopathy and sarcoglycanopathy muscle biopsies showed decreased or absent corresponding proteins on immunohistochemistry and western blotting. FSHD type 1 was diagnosed on the basis of D4Z4 repeat contractions with the 4qA haplotype, and FSHD type 2 was diagnosed by Sanger sequencing, as described previously.^29, 30^

### Pathological evaluation

We routinely performed a battery of histochemical and immunohistochemical staining procedures for diagnostic pathologic evaluation, including hematoxylin and eosin (H&E), modified Gömöri trichrome, acid phosphatase, alkaline phosphatase, cytochrome c oxidase (COX), MxA, Human leukocyte antigen (HLA)-ABC (clone W6/32, Thermo Fisher Scientific), HLA-DR (clone B308, Affinity BioReagents), membrane attack complex (C5b-9, clone aE11 Dako) staining, all of which were re-evaluated for this study. For ASS, pathology slides of the following immunohistochemical stains, which are also routinely performed for diagnosis, were evaluated: neonatal myosin heavy chain (clone WB-MHCn, Leica), utrophin (clone DRP3/20C5, Novocastra), CD3 (polyclonal, Abcam), CD8 (clone DK25, Dako), CD20 (clone L26, eBioscience), and CD68 (clone KP1, Dako). For inclusion body myositis, p62 (SQSTM1; Santa Cruz Biotechnology, Inc.) was also evaluated. The absence or reduction of dysferlin (dysferlin clone Haml/7B6) and sarcoglycans (α-sarcoglycan clone Adl/20A6; ≥-sarcoglycan clone ≥-sarc/5B1; γ-sarcoglycan clone 35DAG/21B5 from Leica Biosystems Newcastle Ltd.) led to the diagnosis of dysferlinopathy and sarcoglycanopathy, respectively.

At the time of pathology slide re-evaluation, muscle histology was blindly scored without autoantibody information. We evaluated four pathology domains (muscle fiber, inflammatory, vascular, and connective tissue) inspired by a pathology scoring system for juvenile dermatomyositis.^31,32^ Although many of the features in the muscle fiber and inflammatory domains used in this study followed the original version of the scoring system, we further modified the evaluation criteria.^16^ For the vascular domain, we compared capillary: myofiber ratio in adult patients with the ratio from twelve histologically normal muscle biopsies as well as among antibody subtypes. We excluded juvenile patients from the capillary: myofiber ratio comparison because limited number of ASS patients in this age group. We categorized the pathological findings of ASS into four patterns: (1) normal/non-specific change, (2) necrotizing myopathy with perifascicular necrosis,^19^ (3) necrotizing myopathy without PFN, and (4) others. The “others” category included but was not limited to the presence of type 2 fiber atrophy, perifascicular atrophy, rimmed vacuoles, nemaline body, cytoplasmic body, and neurogenic change without other distinct pathology. We classified HLA-DR expression into seven categories: 0 = negative, 1 = scattered without a specific pattern, 1+ = patchy to diffusely positive without a specific pattern, 2 = a few positive fibers in the perifascicular area, 3 = several or more positive fibers in the perifascicular area, 4 = positive fibers mainly located in the perifascicular area and involving at least 2/3 of one side of a fascicle, 5 = a mixture of pattern 4 and pattern 1/1+. We grouped HLA-DR categories 3, 4, and 5 as “possible” perifascicular pattern and grouped category 4 and 5 as “perifascicular pattern” since we speculated that these patterns are in continuum (Figure 1A). Other evaluation criteria for histochemical and immunohistochemical staining are described elsewhere.^16,19^ Ultrastructural evaluation for tubuloreticular inclusions (TRI) was performed in 35 available ASS biopsy specimens. Biopsy specimens without TRI observed in 15 randomly examined capillaries were classified as “negative for TRI.”

**Figure 1.**
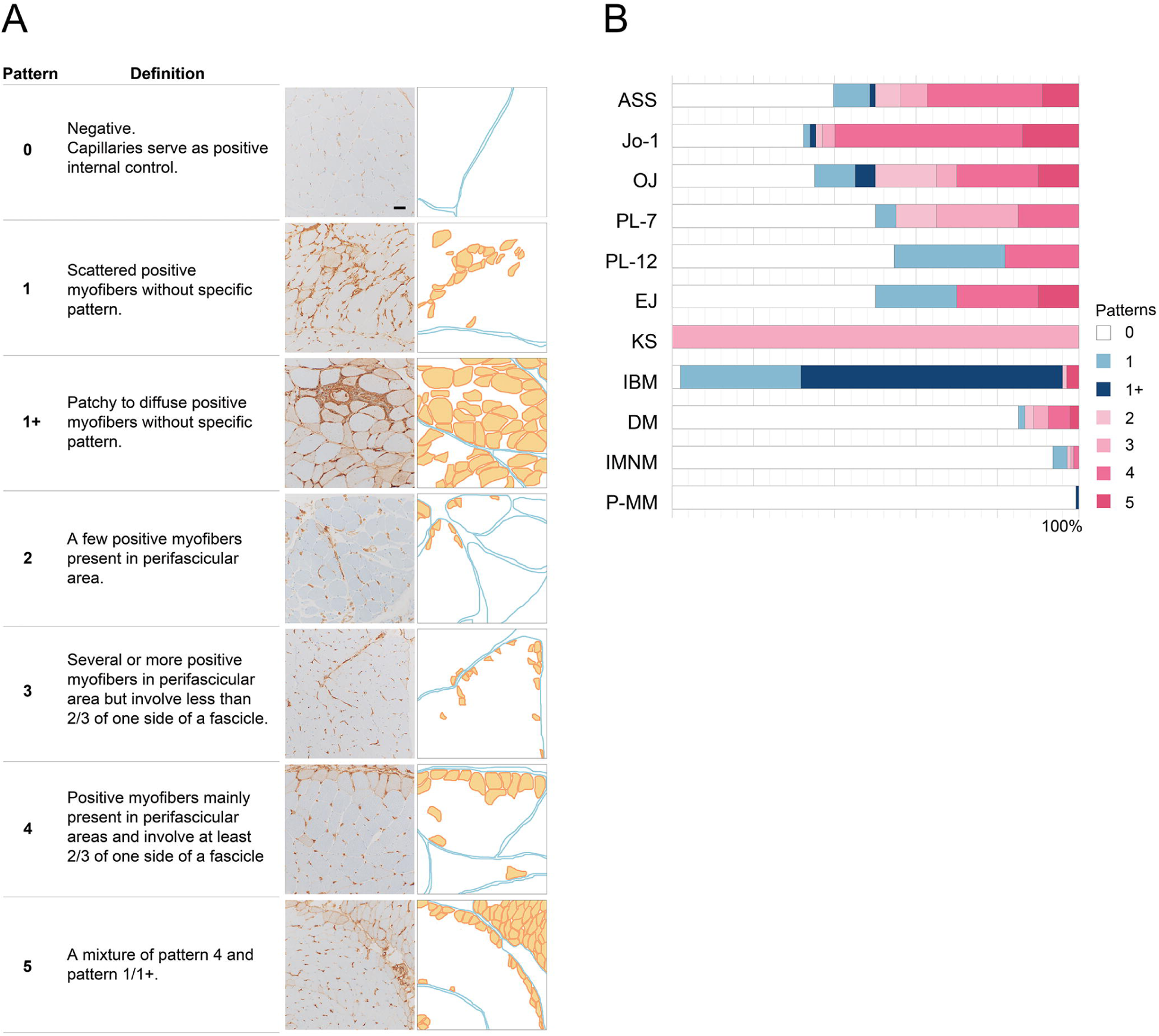
(A) HLA-DR staining patterns. (B) HLA-DR staining pattern in different entities: HLA-DR expression in perifascicular area (pattern 4+5) was more common in ASS than the other idiopathic inflammatory myopathies and potentially myositis mimics. The pattern was more common in anti-Jo-1 ASS than the other antisynthetase syndrome antibodies. **Note:** Bar = 50 μm; HLA-DR, human leukocyte antigen-DR; ASS, antisynthetase syndrome (all subtypes); IBM, inclusion body myositis; DM, dermatomyositis; IMNM, immune-mediated necrotizing myopathy; P-MM, possible myositis mimics.

### Statistical analysis

We included anti-KS and anti-ARS_NOS for the total calculations of patients with ASS but not for the comparisons because of the limited number of patients in the former and the mixed nature of the latter. We used Brown-Forsythe ANOVA followed by Dunnett T3 test for multiple comparisons of the continuous variables. Welch *t* test and Fisher’s exact test were used to compare the continuous and categorical variables, respectively. We compared the percentages of HLA-DR expression and the expression patterns between ASS and non-ASS diseases. The sensitivity, specificity, positive predictive value (PPV), and negative predictive value (NPV) of all HLA-DR expression patterns, including “possible” perifascicular (pattern 3+4+5) and perifascicular (pattern 4+5) patterns were calculated. The clinical and pathological differences between anti-Jo1 ASS and anti-OJ ASS with and without perifascicular HLA-DR expression were compared. Statistical significance was defined by *p* values < 0.05. Statistical analyses were conducted using GraphPad Prism version 9.1.0 for MAC (GraphPad, San Diego, CA, USA).

### RNA sequencing

We performed RNA sequencing analysis using frozen muscle biopsy samples from 152 patients with autoimmune myositis, including 24 patients with ASS (anti-Jo-1 =12 and anti-OJ = 12), 81 with DM (anti-TIF1-γ = 18, anti-Mi-2 = 14, anti-NXP-2 = 32, and anti-MDA5 = 17), 24 with IMNM (anti-SRP = 12 and anti-HMGCR = 12), and 9 with IBM. Twelve histologically normal muscle biopsies were analyzed. In brief, RNA was prepared using TRIzol and Maxwell® RSC Simply RNA Kit (Promega, Madison, WI) and sequenced using the Illumina Hiseq 4000 (Illumina, San Diego, CA) or MGISEQ-2000 (Beijing Genomics Institution, Shenzhen, China). Reads were aligned using Salmon version 0.13.1.^33^ The abundance of each gene was quantified using Salmon version 0.13.1.^33^ Differential gene expression analysis was performed using DESeq2, version 1.34.0. ^34^. Adjusted *p* values (padj) ≤ 0.05 were considered to indicate statistical significance. Differentially expressed genes were ranked according to their degree of significance based on their adjusted *p* value.

### Interferon genes and pathway

The genes listed in the interferon (IFN) signaling pathway were collected from Reactome biological pathways (reactome.org). R version 4.1.3,^35^ RStudio version 2022.02.01,^36^ and the ComplexHeatmap package version 2.10.0^37^ were used to create a differential gene expression heatmap.

### Standard protocol approvals, registrations, and patient consents

The institutional review board of the NCNP approved this study. All materials used in this study were obtained for diagnostic purposes and were permitted for research use with written informed consent.

## Data availability

Anonymized data not published in this article will be made available upon request by any qualified investigator.

## Results

### Clinical features

Among the 212 patients with ASS, 208 (98.1%) were adults and 128 (60.4%) were women. When ASS patients were subclassified by positive anti-ARS antibodies, anti-OJ ASS patients were older (66.2 ± 13.3 vs. 58.3 ± 15.9 years, *p* = 0.03), had higher creatine kinase (CK) levels (6105.6 ± 5724.3 vs. 2930.0 ± 3476.3 U/L, *p* = 0.03), and shorter disease duration before muscle biopsy (10.2 ± 26.3 vs. 29.7 ± 65.1 months, *p* = 0.03) than non-OJ ASS patients. Anti-PL-12 ASS patients had lower CK levels than non-anti-PL-12 ASS patients (1091.6 ± 2146.2 vs. 3651.9 ± 4130.0, *p* = 0.003). Anti-Jo1 ASS was less associated with fever than non-anti-Jo1 ASS, (16.9% vs. 37.1%, *p* = 0.02). The prevalence of muscle weakness, mechanic’s hands, Raynaud phenomenon, joint involvement, ILD, and malignancy did not differ among the ASS subtypes. DM skin lesions, as defined by the 239th European Neuromuscular Center consensus for DM (i.e., presence of Gottron sign/papule and/or heliotrope rash)^9^, were observed in 28.3% of the patients with ASS and were more common in anti-PL7 ASS patients than in non-anti-PL-7 ASS patients (45% vs. 21.5%, p = 0.05) (Table 1, Supplementary eTable 1).

**Table 1.** Clinical features of antisynthetase syndrome in this study.

|  | Anti-ARS antibody |  |  |  |  |  |  |  |
| --- | --- | --- | --- | --- | --- | --- | --- | --- |
|  | All ARS<br>(n = 212) | Jo-1<br>(n = 65) | OJ<br>(n = 20) | PL-7<br>(n = 20) | PL-12<br>(n = 11) | EJ<br>(n = 10) | KS<br>(n = 1) | ARS_NOS<br>(n = 85) |
| Age at biopsy (years) | 59.6±15.9 | 59.1±16.5 | 66.2±13.3* | 55.0±14.8 | 60.5±12.4 | 62.4±11.5 | 13.0 | 59.7±16.2 |
| CK (U/L) | 3100.0±3792.8 | 2742.3±3025.8 | 6105.6±5724.3* | 4484.7 ± 4485.5 | 1091.6 ± 2146.2* | 1866.2±1642.4 | 14900 | 2606.8±3322.7 |
| Disease duration before biopsy (months) | 24.8±55.3<sup>a</sup> | 30.0±70.6<sup>a</sup> | 10.2 ± 26.3* | 17.4 ± 33.0 | 38.2 ± 72.5 | 47.2 ± 773.7 | 5 | 21.9±46.0 |
| Muscle weakness | 197 (92.9) | 61 (93.8) | 20 (100.0) | 19 (95.0) | 10 (90.9) | 10 (100.0) | 1 | 76 (89.4) |
| DM skin lesion<sup>b</sup> | 60 (28.3) | 13 (20.0) | 4 (20.0) | 9 (45.0)* | 3 (27.3) | 2 (20.0) | 1 | 28 (32.9) |
| Mechanic hands | 57 (26.9) | 15 (23.1) | 2 (10.0) | 4 (20.0) | 2 (18.2) | 2 (20.0) | 0 | 32 (37.6) |
| Raynaud's phenomenon | 10 (4.7) | 2 (3.1) | 0 | 0 | 0 | 1 (10) | 0 | 7 (8.2) |
| Joint involvement | 47 (22.2) | 21 (32.3) | 4 (20.0) | 4 (20.0) | 3 (27.3) | 2 (20.0) | 0 | 13 (15.3) |
| Interstitial lung disease | 156 (73.6) | 43 (66.2) | 17 (85.0) | 18 (90.0) | 6 (54.5) | 10 (100.0) | 0 | 61 (71.8) |
| Fever | 57 (26.9) | 11 (16.9)* | 7 (35.0) | 8 (40.0) | 4 (36.4) | 3 (30.0) | 1 | 23 (27.1) |

Continuous data is shown as mean and ± standard deviation while categorical data is reported as number and (percentage).
Abbreviations: ARS, anti-tRNA synthetase; CK, creatine kinase; DM, dermatomyositis; NOS, not otherwise specified.
<sup>a</sup>Information was not available in one case.
<sup>b</sup>DM skin lesions as defined by the 239<sup>th</sup> ENMC workshop include Gottron sign/papule and/or heliotrope rash
\*p < 0.05 compared to the other antibody subtypes

### Ultrastructural study

TRIs were observed in 60% of the ASS muscle biopsies (11/35: anti-Jo-1, 2/10; anti-OJ, 2/5; anti-PL7, 1/3; anti-PL-12, 4/5; anti-EJ, 0/3; and anti-ARS_NOS, 2/9).

### Anti-OJ ASS pathology was prominent in all four domains

Anti-OJ ASS patients had higher muscle fiber domain (4.6 ± 2.0 vs. 2.8 ± 1.8, *p* = 0.001), necrotic fiber (1.7 ± 0.6 vs. 1.0 ± 0.7, *p* < 0.0001), regenerating fiber (0.9 ± 0.4 vs. 0.6 ± 0.5, *p* = 0.008), and perifascicular atrophy scores (0.7 ± 0.9 vs. 0.2 ± 0.6, *p* = 0.04) than non-OJ ASS patients. Anti-Jo-1 ASS patients had lower muscle fiber domain (2.5 ± 1.4 vs. 3.7 ± 2.2, *p* = 0.0003), necrotic fiber (0.9 ± 0.7 vs. 1.3 ± 0.7, *p* = 0.002) atrophic fiber (0.6 ± 0.7 vs. 0.9 ± 0.9, *p* = 0.03), and PFA (0.1 ± 0.4 vs. 0.5 ± 0.8, *p* = 0.0004) scores than non-Jo-1 ASS patients (Figure 2A, Table 2, Supplementary eTable 2). Anti-OJ ASS was associated with a higher inflammatory score (6.8 ± 3.2 vs. 4.5 ± 2.9, *p* = 0.006), endomysial and perimysial CD68 infiltration scores (1.9 ± 0.4 vs. 1.6 ± 0.6, *p* = 0.02 and 1.6 ± 0.7 vs. 1.0 ± 0.8, *p* = 0.0007, respectively), perivascular inflammatory cell infiltration score (0.8 ± 0.4 vs. 0.4 ± 0.5, *p* = 0.002), and a higher prevalence of perivascular inflammatory cell infiltration (75.0% vs. 36.5%, *p* = 0.002) and vasculitis (45.0% vs. 11.2%, *p* = 0.0009) than non-OJ ASS. Anti-PL-12 ASS was associated with lower inflammatory score (2.6 ± 1.5 vs. 5.0 ± 3.1, *p* = 0.0008) (Figure 2B, Table 2, Table 3, and Supplementary eTable 3). For the vascular domain, the capillary:myofiber ratio in adult patients of all ASS subtypes except anti-PL-7 ASS was lower than that of the control specimen (1.1 ± 0.1). The adult capillary:myofiber ratio among antibody subtypes were not distinctively different (Figure 2C, Table 2). Anti-EJ was associated with high muscle domain score than non-EJ ASS (4.7 ± 2.1 vs. 3.0 ± 1.8, *p* = 0.02) but the inflammatory score was not distinctively different from the other subtypes. For the connective tissue domain, anti-OJ and anti-EJ ASS showed more frequent increments in perimysial alkaline phosphatase activity (70.0% vs. 42.1%, *p* = 0.03, and 90.0% vs. non-EJ 43.1%, *p* = 0.06, respectively). Endomysial fibrosis was more common in the anti-EJ ASS group (60.0% vs. 12.1%, p = 0.001). Perimysial alkaline phosphatase activity and endomysial fibrosis were less common in anti-Jo-1 ASS than in non-anti-Jo-1 ASS (33.8% vs. 59.7%, *p* = 0.004 and 7.7% vs. 24.2%, *p* = 0.01, respectively). Perimysial connective tissue fragmentation did not differ among ASS subtypes (Table 2, Figure 3).

**Table 2.** Pathology domains in antisynthetase syndrome.

|  | Anti-ARS antibody |  |  |  |  |  |  |  |
| --- | --- | --- | --- | --- | --- | --- | --- | --- |
|  | All ARS<br>(n = 212) | Jo-1<br>(n = 65) | OJ<br>(n = 20) | PL-7<br>(n = 20) | PL-12<br>(n = 11) | EJ<br>(n = 10) | KS<br>(n = 1) | ARS_NOS<br>(n = 85) |
| **Muscle fiber domain** |  |  |  |  |  |  |  |  |
| Score | 3.0±1.8 | 2.5±1.4* | 4.6±2.0* | 3.1±2.1 | 2.5±1.9 | 4.7±2.1* | 4.0 | 2.8±1.7 |
| **Inflammatory domain** |  |  |  |  |  |  |  |  |
| Score | 4.9±3.0<sup>a</sup> | 4.9±3.2<sup>b</sup> | 6.8±3.2* | 4.2±1.9 | 2.6±1.5<sup>c,*</sup> | 4.5±3.0 | 2.0 | 5.0±3.0<sup>b</sup> |
| **Vascular domain** |  |  |  |  |  |  |  |  |
| Capillary: myofiber ratio | 0.8±0.4<sup>d</sup> | 0.9±0.5<sup>c</sup> | 0.8±0.2<sup>b</sup> | 0.9±0.5<sup>b</sup> | 0.8±0.3 | 0.6±0.3 | 0.8 | 0.8±0.4<sup>e</sup> |
| Adult capillary myofiber ratio<sup>f</sup> | 0.8±0.4<sup>d</sup> | 0.9±0.5<sup>c</sup> | 0.8±0.2<sup>b</sup> | 0.9±0.5<sup>b</sup> | 0.8±0.3 | 0.6±0.3 | 0.8 | 0.8±0.4<sup>e</sup> |
| **Connective tissue domain** |  |  |  |  |  |  |  |  |
| PM-Fr | 109 (51.9)<sup>c</sup> | 29 (44.6) | 14 (70.0) | 11 (55.0) | 5 (45.5) | 7 (60.0) | 0 | 44 (53.0) |
| PM-ALP | 102 (48.1) | 22 (33.8)* | 14 (70.0)* | 10 (50.0) | 3 (27.3) | 9 (90.0)* | 1 | 43 (50.6) |
| Endomysial fibrosis | 34 (16.2)<sup>c</sup> | 5 (7.7)* | 4 (20.0) | 2 (10.0) | 3 (27.3) | 6 (60.0)* | 0 | 14 (16.9) |

Continuous data is shown as mean and ± standard deviation while categorical data is reported as number and (percentage).
Abbreviations: ARS, anti-tRNA synthetase; nos, not otherwise specified; PM-Fr, perimysial fragmentation; PM-ALP, increased perimysial alkaline phosphatase activity; NOS, not otherwise specified.
<sup>a</sup>Four cases were excluded from the analysis due to artifacts
<sup>b</sup>One case was excluded from the analysis due to artifacts
<sup>c</sup>Two cases were excluded from the analysis due to artifacts
<sup>d</sup>Seven cases were excluded from the analysis due to artifacts
<sup>e</sup>Three cases were excluded from the analysis due to artifacts
<sup>f</sup>Four patients were children (<18 years old): Jo-1 = 1, PL-7 = 1, KS = 1, ARS\_NOS = 1
\*p < 0.05 compared to the other antibody subtypes

**Table 3.** Myopathology patterns and histological features of interest in antisynthetase syndrome.

|  | Anti-ARS antibody |  |  |  |  |  |  |  |
| --- | --- | --- | --- | --- | --- | --- | --- | --- |
|  | All ARS<br>(n = 212) | Jo-1<br>(n = 65) | OJ<br>(n = 20) | PL-7<br>(n = 20) | PL-12<br>(n = 11) | EJ<br>(n = 10) | KS<br>(n = 1) | ARS_NOS<br>(n = 85) |
| Myopathology patterns<sup>a</sup> |  |  |  |  |  |  |  |  |
| Normal/non-specific | 37 (17.6) | 13 (20.0) | 0* | 2 (10.0) | 3 (27.3) | 2 (20.0) | 0 | 17 (20.5) |
| Necrotizing myopathy without PFN | 97 (46.2) | 25 (38.5) | 10 (50.0) | 7 (35.0) | 6 (54.5) | 5 (50.0) | 0 | 44 (53.0) |
| Necrotizing myopathy with PFN | 59 (28.1) | 19 (29.2) | 9 (45.0) | 11 (55.0)* | 0* | 3 (30.0) | 1 (100.0) | 16 (19.3) |
| Others | 17 (8.1) | 8 (12.3)<sup>b,c</sup> | 1 (5.0)<sup>d</sup> | 0 | 2 (18.2)<sup>e</sup> | 0 | 0 | 6 (7.2)<sup>f</sup> |
| Histological features of interest |  |  |  |  |  |  |  |  |
| PFN | 60 (28.3) | 20 (30.8) | 9 (45.0) | 11 (55.0)* | 0* | 3 (30.0) | 1 (100.0) | 16 (18.8) |
| PFA | 36 (17.0) | 6 (9.2)* | 8 (40.0)* | 3 (15.0) | 2 (18.2) | 6 (60.0)* | 1 (100.0) | 10 (11.8) |
| PF-COX, decreased | 9 (4.2) | 2 (3.1) | 2 (10.0) | 1 (5.0) | 0 | 1 (9.1) | 0 | 3 (3.5) |
| Vasculitis | 36 (17.0) | 8 (12.3) | 9 (45.0)* | 3 (15.0) | 0 | 1 (9.1) | 0 | 15 (17.6) |

Categorical data is reported as number and (percentage). \*p < 0.05 compared to the other antibody subtypes
Abbreviations: ARS, anti-tRNA synthetase; ACP, acid phosphatase; CD, cluster of differentiation; COX, cytochrome C oxidase; PF, perifascicular; PFA, perifascicular atrophy; PFN, perifascicular necrosis; NOS, not otherwise specified
<sup>a</sup> Two cases of ARS\_NOS were excluded from the analysis due to artifacts
<sup>b</sup> One case classified as granulomatous myositis showed concurrent PFN and granulomatous inflammation
<sup>c</sup> Type 2 fiber atrophy (1), Neurogenic change (4), rimmed vacuole (1), nemaline bodies (1)
<sup>d</sup> Type 2 fiber atrophy (1)
<sup>e</sup> Type 2 fiber atrophy (1), rimmed vacuole (1)
<sup>f</sup> Neurogenic change (2), rimmed vacuole (2), nemaline bodies (1), cytoplasmic body (1)

**Figure 2.**
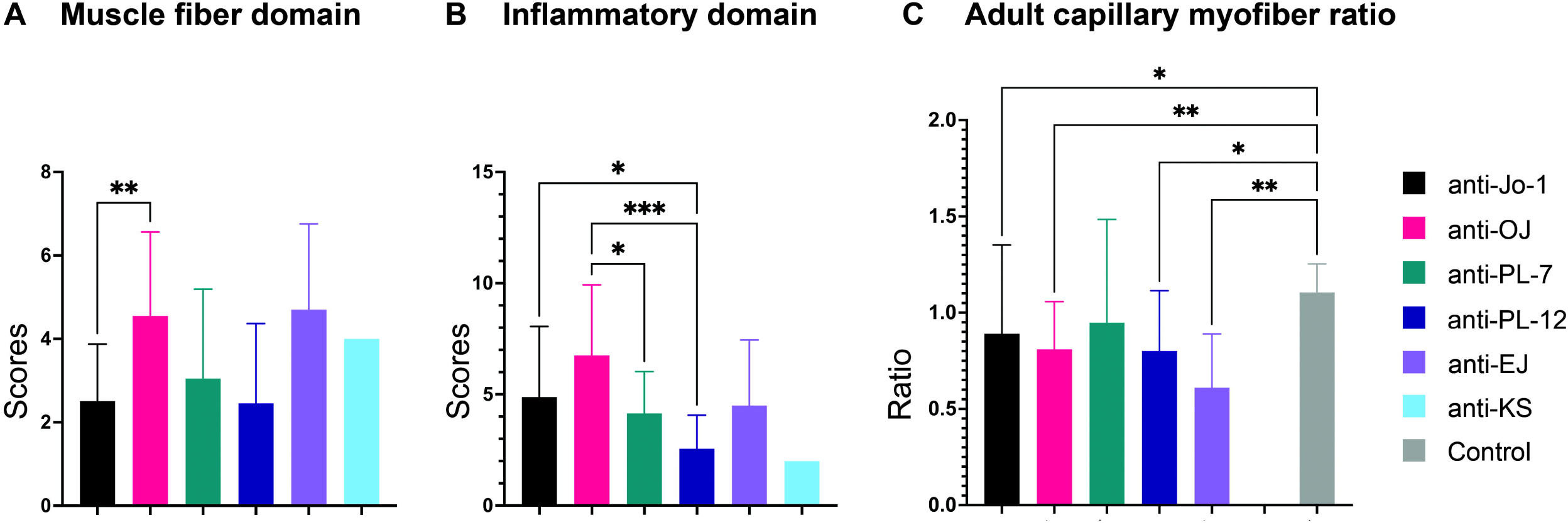
Pathology domains among antisynthetase antibody subtypes. (**A)** Muscle fiber domain: Anti-OJ had a distinctively higher muscle fiber domain score than anti-Jo-1. **(B)** Inflammatory domain: Anti-OJ presented the highest inflammatory domain score which was distinctively higher than anti-PL-7 and anti-PL-12. Anti-Jo-1 showed distinctively higher score than anti-PL-12. **(C)** Adult capillary:myofiber ratio: The ratio in all antisynthetase antibody subtypes was lower than controls. The ratio was not distinctively different among antibody subtypes. Bar = mean±SD, analysis of variance with Dunnett T3 multiple comparison: *p* value 0.0332 (*), 0.0021 (**), 0.0002(***), < 0.0001(****).

**Figure 3.**
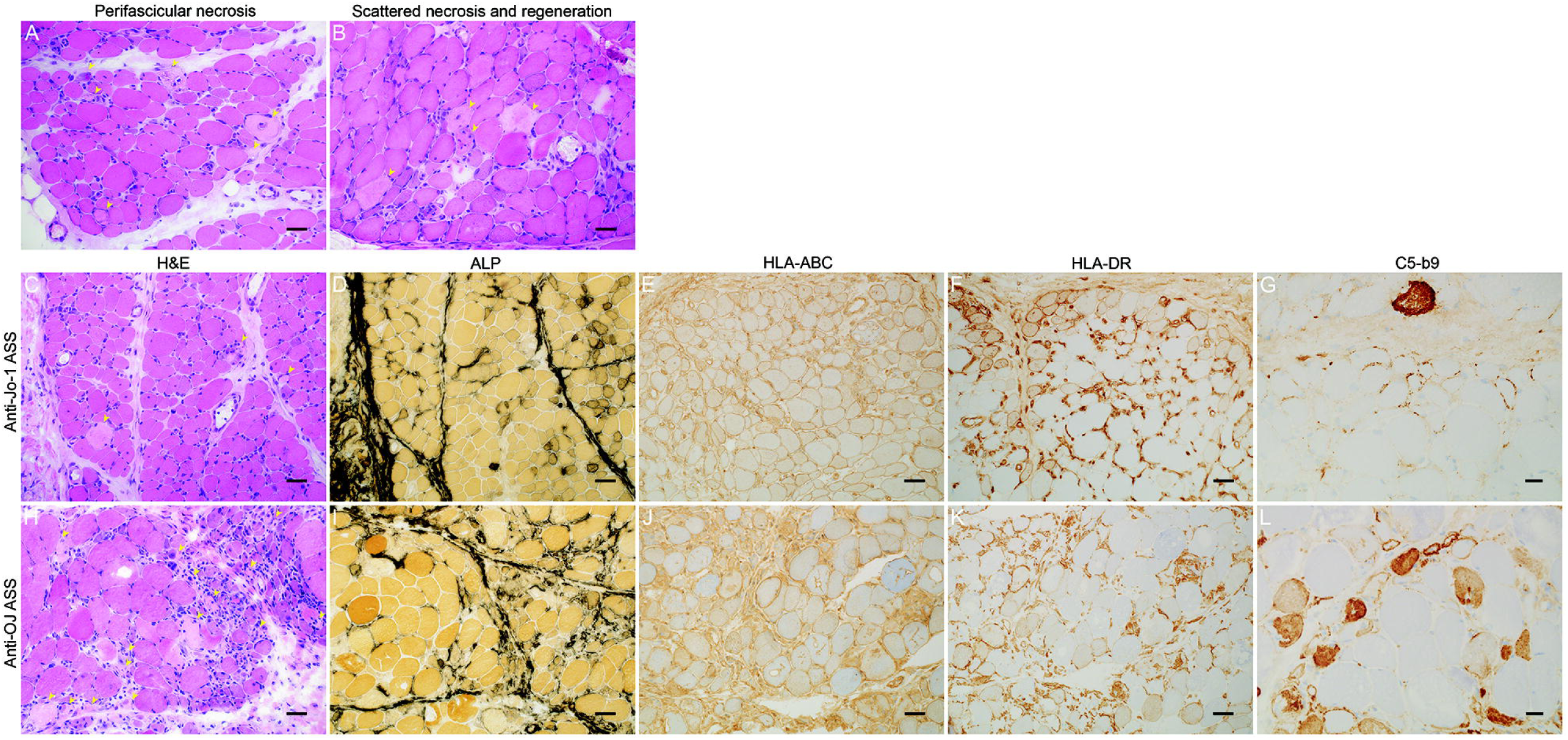
Histologic patterns in antisynthetase syndrome. **(A)** Perifascicular necrosis: more than 2/3 of necrotic fibers (yellow arrowheads) are present at the perifascicular areas. Perimysial connective tissue fragmentation is also observed. **(B)** Scattered necrotic and regenerating fibers without specific pattern. Representative figures for anti-Jo-1 ASS **(C-G)** and anti-OJ ASS **(H-L):** Both anti-Jo-1 and anti-OJ show perifascicular necrosis on H&E **(C and H)**, but more prominent inflammatory cell infiltration is observed in anti-OJ ASS **(H)**. Both anti-Jo-1 and anti-OJ ASS show increased perimysium alkaline phosphatase activity **(D and I)**, diffuse and strong HLA-ABC positivity **(E and J)** and sarcolemmal C5b-9 expression in perifascicular areas **(G and L)**. In this figure, anti-Jo-1 ASS shows perifascicular HLA-DR positivity **(F)** while anti-OJ ASS shows scattered positivity **(K) Note:** A-B, C-F, H-K bars = 50 μm; G-L bars = 20 μm; H&E, hematoxylin and eosin; ALP, alkaline phosphatase; HLA-ABC, human leukocyte antigen-ABC; HLA-DR, human leukocyte antigen-DR; C5b-9: membrane attack complex; yellow arrowheads in A-C and H = necrotic fibers

### Necrotizing myopathy was the most common myopathological pattern in ASS

The most common myopathological pattern in ASS patients was necrotizing myopathy without perifascicular necrosis (46.2%), and the prevalence of this pattern did not differ substantially among the ASS subtypes. Perifascicular necrosis was present in 28.3% of the overall ASS cases and was more common in anti-PL-7 ASS (55.0% vs. non-PL-7 30.8%, *p* = 0.04). Perifascicular necrosis was not present in the anti-PL-12 ASS group (0% vs. non-PL-12 37.9%, *p* = 0.008). Perifascicular atrophy was present in 17% of all ASS cases; it was common in anti-OJ (40.0% vs. non-OJ 16.8%, *p* = 0.03) and anti-EJ ASS (60.0% vs. 17.1%, *p* = 0.005). Decreased COX activity in perifascicular fibers, CD8 or CD68/acid phosphatase-positive cell infiltration into non-necrotic fibers, and CD20-positive cell aggregation were not common in ASS and were not distinctively different among ASS subtypes (Table 3 and Supplementary eTable 4).

### Perifascicular HLA-ABC enhancement and HLA-DR localization was common in anti-Jo-1 ASS

HLA-ABC expression was present in 92.5% of ASS muscle biopsies, and HLA-ABC expression with perifascicular enhancement was more common in anti-Jo-1 ASS (26.2% vs. non-Jo-1 3.2%, *p* = 0.0003). Necrotic and regenerating fibers were present in only 1 of 16 HLA-ABC-negative cases. HLA-DR expression was observed in 60.4% of ASS patients. All but one HLA-DR-positive case showed HLA-ABC positivity. The most common HLA-DR expression pattern was perifascicular localization (pattern 4) (28.3%), which was distinctively common in anti-Jo1 ASS (46.2% vs. non-Jo-1 17.7%, *p* = 0.0007) (Figure 1B, Table 4, Supplementary eTable 5). Pattern 4 HLA-DR expression was observed in 40.7% (24/59) of ASS muscle biopsies showing necrotizing myopathy with perifascicular necrosis, 29.9% (29/97) of necrotizing myopathy without perifascicular necrosis, 16.2% (6/37) of normal/nonspecific pathology, and one muscle biopsy with neurogenic changes (Table 3, Supplementary eTable 6). Membrane attack complex deposition on capillaries and sarcolemma was noted in 58.8% and 47.2% of all ASS cases, respectively. Sarcoplasmic MxA expression was observed in three ASS patients (1.4%, 1 Jo-1, 1 OJ, and 1 PL-7-positive patient)^21^; all of them were HLA-DR-negative. The expressions of membrane attack complex and MxA were not distinctively different among the ASS subtypes (Table 4).

**Table 4.** Immunohistochemical feature in antisynthetase syndrome.

|  | Anti-ARS antibody |  |  |  |  |  |  |  |
| --- | --- | --- | --- | --- | --- | --- | --- | --- |
|  | All ARS<br>(n = 212) | Jo-1<br>(n = 65) | OJ<br>(n = 20) | PL-7<br>(n = 20) | PL-12<br>(n = 11) | EJ<br>(n = 10) | KS<br>(n = 1) | ARS_NOS<br>(n = 85) |
| HLA-ABC expression | 196 (92.5) | 61 (93.8) | 20 (100.0) | 20 (100.0) | 9 (81.8) | 10 (100.0) | 1 (100.0) | 75 (88.2) |
| HLA-ABC expression with PF enhancement | 36 (17.0) | 17 (26.2)* | 1 (5.0) | 0 | 0 | 1 (10.0) | 0 | 17 (20.0) |
| HLA-DR expression | 128 (60.4) | 44 (67.7) | 13 (65.0) | 10 (50.0) | 5 (45.5) | 5 (50.0) | 1 (100.0) | 50 (58.8) |
| MAC capillary deposition | 124 (58.8)<sup>a</sup> | 42 (64.6) | 10 (50.0) | 11 (65.0) | 6 (54.5) | 4 (45.0) | 0 | 49 (58.3)<sup>a</sup> |
| MAC capillary deposition in PF area | 13 (6.2)<sup>a</sup> | 6 (9.2) | 1 (5.0) | 0 | 2 (18.2) | 0 | 0 | 4 (4.8) |
| MAC sarcolemmal deposition | 100 (47.2) | 27 (41.5) | 10 (50.0) | 8 (40.0) | 2 (18.2) | 8 (80.0)* | 0 | 45 (52.9) |
| MAC sarcolemmal deposition in PF area | 64 (30.2) | 19 (29.2) | 7 (35.0) | 4 (20.0) | 1 (9.1) | 6 (60.0) | 0 | 27 (31.8) |
| MxA expression | 3 (1.4) | 1 (1.5) | 1 (5.0) | 1 (5.0) | 0 | 0 | 0 | 0 |

Categorical data is reported as number and (percentage).
Abbreviations: ARS, anti-tRNA synthetase; HLA, human leukocyte antigen; NOS, not otherwise specified; MAC membrane attack complex (C5b-9); MxA, Myxovirus resistance protein A; PF, perifascicular
<sup>a</sup>One case was not included due to artifacts
\*p < 0.05 compared to the other antibody subtypes

### HLA-DR expression was more common in IBM with patchy/diffuse and scattered patterns

We reviewed HLA-DR expression patterns in other autoimmune myositis subtypes and in P-MM. Among the non-ASS cases, HLA-DR expression was much more common in IBM (98.0% vs. 60.4% in ASS, *p* < 0.0001), with prominent patchy/diffuse (64.4%) followed by scattered (29.7%) patterns. Except for IBM, HLA-DR expression was distinctively more common in ASS than in the other entities (14.9% DM, 6.4% IMNM, 0.7% P-MM, p < 0.0001) (Figure 1B, Supplementary eTable 7).

### Perifascicular HLA-DR expression was highly specific for anti-Jo-1 ASS

The presence of HLA-DR expression in any pattern suggested the diagnosis of ASS over other forms of autoimmune myositis and P-MM with 80.1% specificity, 60.4% sensitivity, 46.4% PPV, and 87.6% and PPV. Excluding 191 MxA-positive muscle biopsies (188 DM and 3 MxA-positive ASS) and 101 muscle biopsies from clinicopathologically diagnosed IBM, HLA-DR expression of any pattern suggested a diagnosis of ASS with 95.4% specificity, 61.2% sensitivity, 85.9% PPV and 84.2% NPV. The presence of perifascicular HLA-DR expression (pattern 4+5) indicated a diagnosis of anti-Jo-1 ASS over other entities (96.5% specificity, 60.9% sensitivity, 68.4% PPV, and 95.2% NPV) (Supplementary eTable 8).

### IFN-γ inducible genes were the most significantly upregulated IFN genes in ASS and IBM

The expression levels of IFN signaling pathway-associated genes differed among autoimmune myositis subtypes. The most significantly upregulated IFN genes in anti-Jo-1, anti-OJ ASS, and inclusion body myositis were the type II IFN (IFN-γ)-inducible genes (e.g. *PSMB8* and *B2M*). Type I IFN-inducible genes (e.g., *ISG15, IFI6, OAS* gene families) were highly upregulated in anti-TIF1-γ, Mi-2, MDA5, and NXP-2 DM. Anti-HMGCR and anti-SRP IMNM showed lower expression of these genes. Although the class II major histocompatibility complex transactivator (*CIITA*) was not present among the 10 most significantly upregulated IFN genes, its expression level in ASS and IBM showed a higher degree of significance than that of the other entities (Figure 4, Supplementary eTable 9).

**Figure 4.**
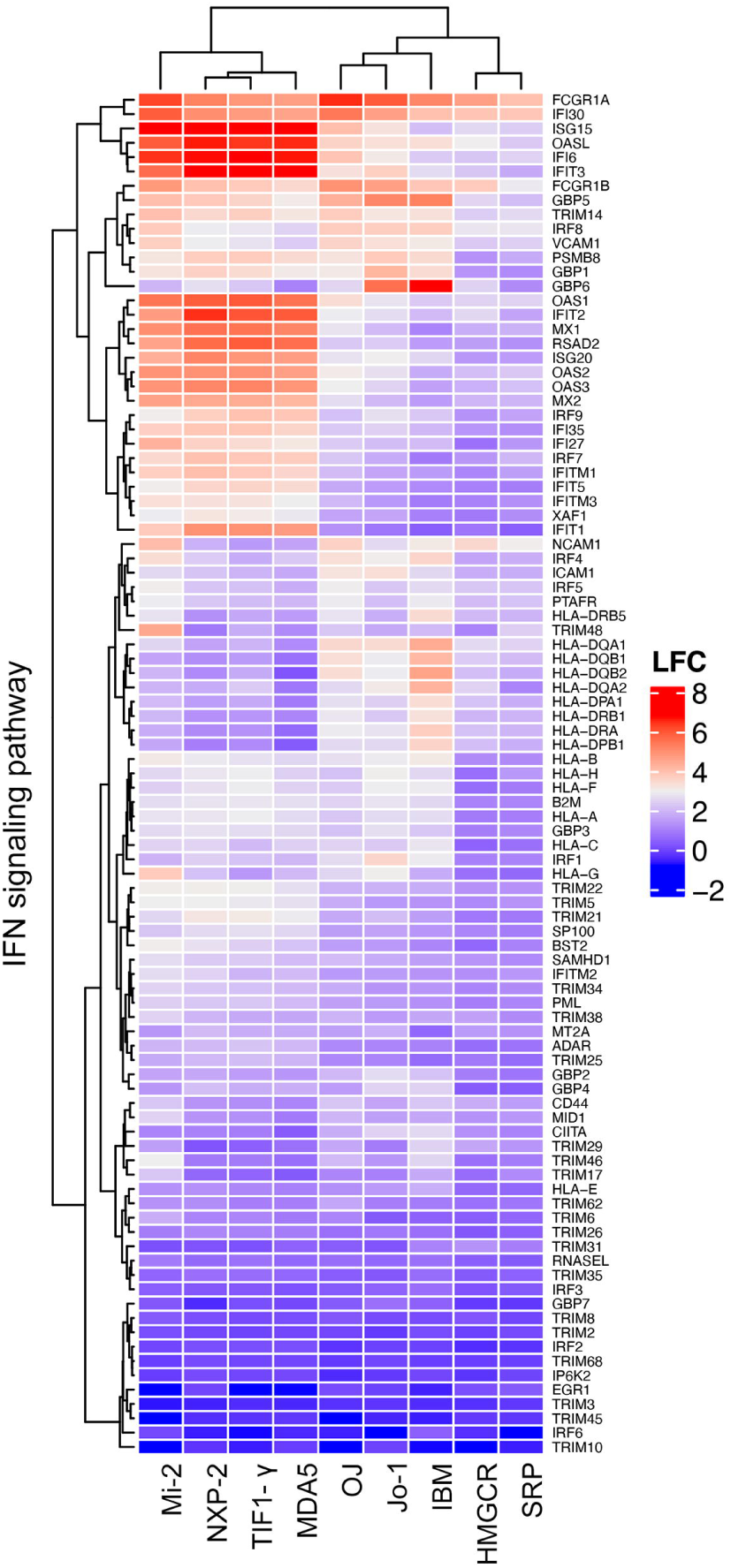
Expression levels of genes in IFN-signaling pathway in different subtypes of idiopathic inflammatory myopathy. **Note:** IFN, interferon; Mi-2, anti-nucleosome remodeling-deacetylase (NuRD) complex; NXP-2, anti-nuclear complex protein 2; TIF1-γ, anti-transcription intermediary factor 1-gamma; MDA5, anti-melanoma differentiation-associated gene 5; OJ, anti-OJ; Jo-1, anti-Jo-1; HMGCR, anti-3-hydroxy-3-methylglutaryl-coenzyme A reductase; SRP, anti-signal recognition particle. LFC = log2 fold change

## Discussion

This was a comprehensive study on ASS myopathology aimed at describing the features of different ASS subtypes, especially non-Jo-1 ASS. Recognition of non-Jo-1 anti-ARS-associated myopathology has been limited not only because of the rarity of the antibodies, but also because of the technical challenges in antibody detection by commercially available kits.^22,38,39^ Notably, line/dot blot immunoassays and ELISA likely fail to detect anti-OJ antibodies due to the structural complexity in recognition of autoantigens in a multi-enzyme synthetase complex.^38^ Immunoprecipitation methods are currently recommended for anti-OJ detection.^22,39^ Thus, the number of anti-OJ ASS cases reported in the literature may be underestimated unless the original method, RNA or protein immunoprecipitation, is applied.^22,38,39^ Notably, all anti-OJ-positive patients in this study were identified by RNA immunoprecipitation.

Anti-OJ ASS showed clinical and myopathological features suggestive of more severe muscle involvement, including older patient age, higher CK level, shorter disease duration before biopsy, higher muscle fiber domain, and inflammatory domain scores. Although the study results could have been partly affected by antibody test bias of seven anti-OJ cases, the overall results conformed with the results of the bias-free original cohort^20-22^ (Supplementary eTable 10, eTable 11). Anti-PL-12 ASS showed features suggestive of mild muscle disease involvement, with lower CK levels and inflammatory domain scores. Our findings for anti-PL-12 ASS support a previous clinical study in which anti-PL-12 ASS showed less muscle involvement at the onset of the disease.^11^ We did not find any difference in extramuscular involvement among the ASS subtypes, except for a lower number of anti-Jo-1 patients experiencing fever before muscle biopsy diagnosis.

Anti-Jo-1 ASS has been categorized as an immune myopathy with perimysial pathology (IMPP) showing a combination of myofiber necrosis in the perifascicular region and perimysial pathology (i.e., perimysial connective tissue fragmentation and increased perimysial alkaline phosphatase activity).^3,40,41^ The presence of perifascicular necrosis, defined by the presence of 2/3 of the total necrotic fiber in the perifascicular region, has been reported in 47%-79% of the cases of anti-Jo-1 ASS.^16,19,20^ However, these features are not exclusively limited to anti-Jo-1 ASS but are also present in anti-Mi-2 DM and other ASS subtypes.^16,20^ The fact that perifascicular necrosis is more commonly present in anti-PL-7 ASS could be explained by the severity of myopathology, which is between the lower end of the spectrum, represented by anti-PL-12 and anti-Jo-1 ASS, and the higher end of the spectrum, represented by anti-OJ and anti-EJ ASS. In parallel with the reduced muscle involvement, anti-PL-12 muscle biopsies did not show perifascicular necrosis. Necrotizing myopathy without perifascicular necrosis was the most common myopathology pattern in ASS. Perimysial connective tissue fragmentation is not distinctively different among ASS subtypes. Anti-OJ and anti-EJ ASS were more commonly associated with perimysial alkaline phosphatase activity. The more frequent perimysial alkaline phosphatase activity in these two entities was likely associated with their high perimysial CD68-positive cell infiltration scores (Supplementary eTable 3), although the scores in anti-EJ ASS and non-EJ ASS were not distinctively different. We speculate that tissue-nonspecific ALPs could be induced by a subset of tumor necrosis factor-α and interleukin-1β-secreting CD68-positive cells.^42,43^ Interestingly, the anti-Jo-1 ASS in our study was less associated with perimysial alkaline phosphatase activity than the non-Jo-1 ASS. A decreased capillary:myofiber ratio has been previously reported for anti-Jo1 ASS.^19.^ The overall reduction in capillary:myofiber ratio in ASS could be caused by vascular damage due to capillary membrane attack complex deposition. However, the indistinctively different capillary:myofiber ratios in anti-PL-7 in comparison with control suggest that currently unidentified additional factor(s) cause different degrees of tissue hypoxia in ASS subtypes. Anti-EJ and anti-OJ ASS are more commonly associated with perifascicular atrophy. This finding could be attributed to the more severe muscle tissue damage in both subtypes but probably due to different pathomechanism considering higher inflammatory domain score in anti-OJ ASS. Whether the underlying pathomechanisms of perimysial alkaline phosphatase activity, decreased capillary:myofiber ratio, and perifascicular atrophy in ASS are different from those in other entities, such as DM showing the same features, remains to be elucidated.

A few smaller studies have addressed the HLA-ABC, HLA-DR, and membrane attack complex findings in ASS.^12,19^ We confirmed that HLA-ABC was present in the majority of ASS cases. Perifascicular HLA-ABC enhancement is more common in anti-Jo-1 than in non-anti-Jo-1 ASS, but at a lower percentage (26.2%) than in a previous study (79%).^19^ HLA-DR myofiber expression was previously reported in 82.8% of the patients in a study involving anti-Jo-1, anti-PL-7, anti-PL-12, and anti-EJ ASS; the findings of that study showed perifascicular HLA-DR distribution in all cases.^17^ Similar findings were observed in a study that focused on anti-Jo-1 ASS.^19^ Our study confirmed the preferential perifascicular HLA-DR myofiber localization in anti-Jo-1 ASS, but at a lower percentage (60.0%). In addition, we observed other HLA-DR staining patterns in all ASS subtypes (Figure 1B).

Clinicopathological exclusion of entities with possible histological and immunohistochemical features mimicking ASS (i.e., exclusion of DM on the basis of MxA positivity and IBM on the basis of clinicopathological criteria) increased the specificity of HLA-DR expression in ASS from 80.1% to 95.4%. The presence of perifascicular HLA-DR expression in this setting was highly specific (96.5%) to anti-Jo-1 ASS. Co-expression of CIITA and HLA-DR on perifascicular fibers reflects activation of the IFN-γ signaling pathway^44, 45^ (Supplementary eFigure 1). Indeed, IFN-γ-related genes, including *CIITA*, are upregulated in IBM myositis and ASS than in other autoimmune myositis subtypes^46^ (Supplementary Table 9). The presence of CD8+ T cells in the vicinity of perifascicular HLA-DR expression in a previous study indicates their role in IFN−γ production.^12^ However, perifascicular HLA-DR expression was also observed in six ASS cases with normal/nonspecific pathology, suggesting that the expression was likely induced by high levels of serum IFN-γ.^47^ Unlike DM, in which TRIs are ubiquitous (100%), these inclusions are present in subpopulations of ASS (47%-60%)^19,48^. Since TRIs can be induced in cultured endothelial cells by type I IFN (IFN1: IFN-α and IFN-β)^49^, the number of TRIs in ASS can also be explained by a lower expression level of genes involved in the IFN1 pathway. We confirmed that capillary membrane attack complex deposition can be present, and that strong positivity in the perifascicular area is uncommon in ASS.^19^ Sarcolemmal membrane attack complex deposition, both in general and in perifascicular areas, is more common in ASS than in DM (47.2% vs. 26.6% and 30.2% vs. 12.5%, respectively).^48^

Due to the retrospective nature of the study, the clinical information recorded was limited and did not represent the prevalence of muscular and extramuscular involvement throughout the disease course. All patients in this study underwent muscle biopsy for diagnostic purposes, suggesting that our cohort may be biased, omitting patients without obvious muscle involvement. We could not comment on a single case of anti-KS. Anti-Ha and anti-Zo antibodies were not detected in this cohort. We included all cases of the entities categorized as P-MM regardless of the pathological pattern, to evaluate myofiber HLA-DR expression. However, only one case expressed HLA-DR. The association of the presence of P-MM pathology in these entities with HLA-DR expression requires larger cohort studies.

Although ASS is serologically categorized as autoimmune myositis^18^, integration of clinical information and pathological profiles, especially the expression of MxA and HLA-DR, can help differentiate ASS from other forms of autoimmune myositis even when serological information is limited. In addition, pathological features can help indicate anti-OJ ASS, even with negative results in the anti-ARS antibody assays obtained from commercial laboratories. The correlation of high serum IFN-γ levels with ASS with myofiber HLA-DR expression and the potential benefits of IFN-γ pathway inhibition for these patients remain to be determined.

## Abbreviations

ACP: Acid phosphatase
ALP: Alkaline phosphatase
ANO5: Anoctamine 5
ASS: Antisynthetase syndrome
ARS: Anti-tRNA synthetase
CD: cluster of differentiation
CK: Creatine kinase
COX: Cytochrome C oxidase
DM: Dermatomyositis
FKRP: Fukutin-related protein
FSHD: facioscapulohumeral muscular dystrophy
H&E: Hematoxylin and eosin
HLA: Human leukocyte antigen
IBM: Inclusion body myositis
IFN: Interferon
IHC: immunohistochemical study
IMNM: Immune mediated necrotizing myopathy
IMPP: Immune myopathy with perimysial pathology
ILD: Interstitial lung disease
MSA: Myositis specific antibody
MDA5: Melanoma differentiation-associated protein 5
MxA: Myxovirus resistance protein A
NCNP: National Center of Neurology and Psychiatry
NPV: Negative predictive value
NOS: Not otherwise specified
PFA: Perifascicular atrophy
PFN: Perifascicular necrosis
PM: Polymyositis
P-MM: Possible myositis mimics
PM-ALP: perimysial alkaline phosphatase activity
PPV: Positive predictive value
TRI: Tubuloreticular inclusion

## Acknowledgements

The authors thank Kazu Iwasawa, Kaoru Tatezawa, Naho Fushimi, Miyuki Matsuda, Hisayoshi Nakamura, and Kousuke Akai for their technical assistance.

## Supplementary material

Supplementary material is available at *Brain* online.

