## Supplementary for "Muscle pathology of antisynthetase syndrome according to antibody subtypes"

**Supplementary eTable 1 Additional clinical features of antisynthetase syndrome in this study**

| | Anti-ARS antibody | | | | | | | |
|---|---|---|---|---|---|---|---|---|
| | **All ARS** (n = 212) | **Jo-1** (n = 65) | **OJ** (n = 20) | **PL-7** (n = 20) | **PL-12** (n = 11) | **EJ** (n = 10) | **KS** (n = 1) | **ARS_NOS** (n = 85) |
| Adult (≥ 18 years old) | 208 (98.1) | 64 (98.5) | 20 (100.0) | 19 (95.0) | 11(100.0) | 10 (100.0) | 0 | 84 (98.8) |
| Woman | 128 (60.4) | 40 (61.5) | 9 (45.0) | 11 (55.0) | 5 (45.5) | 6 (60.0) | 1 (100.0) | 56 (65.9) |
| Immunotherapy at the time of biopsy | 53 (25.0) | 12 (18.5) | 6 (30.0) | 5 (25.0) | 2 (18.2) | 1 (10.0) | 0 | 27 (31.8) |
| Malignancy | 17 (8.0) | 6 (9.2) | 3 (15.0) | 0 | 1 (9.1) | 0 | 0 | 7 (8.2) |

Continuous data is shown as mean and ± standard deviation while categorial data is reported as number and (percentage).
Abbreviations: ARS, anti-tRNA synthetase; NOS, not otherwise specified.

**Supplementary eTable 2 Muscle fiber domain scores in antisynthetase syndrome**

| | Anti-ARS antibody | | | | | | | |
|---|---|---|---|---|---|---|---|---|
| | **All ARS** (n = 212) | **Jo-1** (n = 65) | **OJ** (n = 20) | **PL-7** (n = 20) | **PL-12** (n = 11) | **EJ** (n = 10) | **KS** (n = 1) | **ARS_NOS** (n = 85) |
| **Muscle fiber domain** | | | | | | | | |
| Total Score | 3.0±1.8 | 2.5±1.4* | 4.6±2.0* | 3.1±2.1 | 2.5±1.9 | 4.7±2.1* | 4.0 | 2.8±1.7 |
| Necrotic fiber score | 1.1±0.8 | 0.9±0.7* | 1.7±0.6* | 1.3±0.6 | 0.7±0.8 | 1.3±0.8 | 2.0 | 2.5±1.4 |
| Regenerating fiber score | 0.6±0.5 | 0.6±0.5 | 0.9±0.4* | 0.6±0.5 | 0.4±0.5 | 0.8±0.4 | 1.0 | 0.6±0.5 |
| Atrophic fiber score | 0.7±0.8 | 0.6±0.7* | 1.0±0.9 | 0.7±0.9 | 0.8±0.9 | 1.2±0.8 | 0 | 0.7±0.8 |
| PFA score | 0.2±0.6 | 0.1±0.4* | 0.7±0.9* | 0.3±0.7 | 0.3±0.6 | 0.8±0.8 | 1.0 | 0.2±0.5 |
| Fiber with internalized nuclei score | 0.3±0.5 | 0.3±0.5 | 0.3±0.5 | 0.2±0.4 | 0.3±0.5 | 0.6±0.5 | 0 | 0.2±0.4 |
| Fiber with internalized nuclei >3% | 60 (28.3) | 22 (33.8) | 6 (30.0) | 4 (21.1) | 3 (27.3) | 6 (54.5) | 0 | 19 (22.4) |

Continuous data is shown as mean and ± standard deviation while categorial data is reported as number and (percentage).
Abbreviations: ARS, anti-tRNA synthetase; NOS, not otherwise specified; PFA, perifascicular atrophy
*p < 0.05 compared to the other antibody subtypes

**Supplementary eTable 3 Inflammatory domain in antisynthetase syndrome**

| | Anti-ARS antibody | | | | | | | |
|---|---|---|---|---|---|---|---|---|
| | **All ARS** (n = 212) | **Jo-1** (n = 65) | **OJ** (n = 20) | **PL-7** (n = 20) | **PL-12** (n = 11) | **EJ** (n = 10) | **KS** (n = 1) | **ARS_NOS** (n = 85) |
| **Inflammatory domain** | | | | | | | | |
| Total Score | 4.9±3.0[a] | 4.9±3.2[b] | 6.8±3.2* | 4.2±1.9 | 2.6±1.5[c,*] | 4.5±3.0 | 2.0 | 5.0±3.0[b] |
| Endomysial CD3 infiltration | 0.7±0.8[a] | 0.8±0.8[b] | 1.1±0.9 | 0.8±0.5 | 0.1±0.3[c,*] | 0.7±0.8 | 0 | 0.7±0.7[b] |
| Perimysial CD3 infiltration | 0.3±0.6[a] | 0.3±0.7[b] | 0.6±0.7 | 0* | 0[c,*] | 0.2±0.4 | 0 | 0.3±0.6[b] |
| Endomysial CD20 infiltration | 0.4±0.7[a] | 0.5±0.7[b] | 0.7±0.7 | 0.4±0.6 | 0.1±0.3[c,*] | 0.2±0.4 | 0 | 0.4±0.7[b] |
| Perimysial CD20 infiltration | 0.2±0.5[a] | 0.2±0.5[b] | 0.3±0.6 | 0.2±0.4 | 0[c,*] | 0.1±0.3 | 0 | 0.3±0.6[b] |
| Endomysial CD68 infiltration | 1.6±0.5[a] | 1.6±0.6[b] | 1.9±0.4* | 1.7±0.5 | 1.3±0.7[c] | 1.7±0.7 | 0 | 1.6±0.5[b] |
| Perimysial CD68 infiltration | 1.1±0.8[a] | 1.0±0.8[b] | 1.6±0.7* | 0.8±0.8 | 0.8±0.7[c] | 1.2±0.9 | 0 | 1.3±0.7[b] |
| Perivascular inflammatory cell infiltration score | 0.4±0.5 | 0.4±0.5 | 0.8±0.4* | 0.4±0.5 | 0.2±0.4 | 0.4±0.5 | 0 | 0.4±0.5 |
| Perivascular inflammatory cell infiltration | 87 (41.0) | 26 (40.0) | 15 (75.0)* | 7 (35.0) | 2 (18.2) | 4 (40.0) | 0 | 33 (38.8) |

Categorial data is reported as number and (percentage).
Abbreviations: ARS, anti-tRNA synthetase; nos, not otherwise specified; PM-Fr, perimysial fragmentation; PM-ALP, increased perimysial alkaline phosphatase activity; NOS, not otherwise specified.
[a]Four cases were excluded from the analysis due to artifacts
[b]One case was excluded from the analysis due to artifacts
[c]Two cases were excluded from the analysis due to artifacts
*$p$ < 0.05 compared to the other antibody subtypes

**Supplementary eTable 4 Additional histological features of interest**

| | Anti-ARS antibody | | | | | | | |
|---|---|---|---|---|---|---|---|---|
| | **All ARS** (n = 212) | **Jo-1** (n = 65) | **OJ** (n = 20) | **PL-7** (n = 20) | **PL-12** (n = 11) | **EJ** (n = 10) | **KS** (n = 1) | **ARS_NOS** (n = 85) |
| CD8 infiltration in non-necrotic fiber | 1 (0.5) | 0 | 0 | 0 | 0 | 0 | 0 | 1 (1.2) |
| CD68/ACP infiltration in non-necrotic fiber | 7 (3.3) | 2 (3.1) | 1 (5.0) | 1 (5.3) | 0 | 1 (9.1) | 0 | 2 (2.4) |
| CD20 aggregation | 10 (4.7) | 4 (6.2) | 1 (5.0) | 1(5.3) | 0 | 0 | 0 | 4 (4.7) |

Categorial data is reported as number and (percentage).
Abbreviations: ARS, anti-tRNA synthetase; ACP, acid phosphatase; CD, cluster of differentiation; NOS, not otherwise specified
*$p < 0.05$ compared to the other antibody subtypes

**Supplementary eTable 5 HLA-DR expression patterns in ASS**

| | Anti-ARS antibody | | | | | | | |
|---|---|---|---|---|---|---|---|---|
| | **All ARS** (n = 212) | **Jo-1** (n = 65) | **OJ** (n = 20) | **PL-7** (n = 20) | **PL-12** (n = 11) | **EJ** (n = 10) | **KS** (n = 1) | **ARS_NOS** (n = 85) |
| HLA-DR expression | 128 (60.4) | 44 (67.7) | 13 (65.0) | 10 (50.0) | 5 (45.5) | 5 (50.0) | 1 (100.0) | 50 (58.8) |
| Pattern 1 | 19 (9.0) | 1 (1.5)* | 2 (10.0) | 1 (5.0) | 3 (27.3)* | 2 (20.0) | 0 | 10 (11.8) |
| Pattern 1+ | 3 (1.4) | 1 (1.5) | 1 (5.0) | 0 | 0 | 0 | 0 | 1 (1.2) |
| Pattern 2 | 13 (6.1) | 1 (1.5)* | 3 (15.0) | 2 (10.0) | 0 | 0 | 0 | 7 (8.2) |
| Pattern 3 | 14 (6.6) | 2 (3.1) | 1 (5.0) | 4 (20.0)* | 0 | 0 | 1 (100.0) | 6 (7.1) |
| Pattern 4 | 60 (28.3) | 30 (46.2)* | 4 (20.0) | 3 (15.0) | 2 (18.2) | 2 (10.0) | 0 | 19 (22.4) |
| Pattern 5 | 19 (9.0) | 9 (13.8) | 2 (10.0) | 0 | 0 | 1 (10.0) | 0 | 7 (8.2) |
| "Possible" perifascicular pattern (3+4+5) | 93 (43.9) | 41 (63.1)* | 7 (35.0) | 7 (35.0) | 2 (18.2) | 3 (30.0) | 1 (100.0) | 32 (37.6) |
| Perifascicular pattern (4+5) | 79 (37.3) | 39 (60.0)* | 6 (30.0) | 3 (15.0)* | 2 (18.2) | 3 (30.0) | 0 | 26 (30.6) |

Categorial data is reported as number and (percentage).
Abbreviations: ARS, anti-tRNA synthetase; HLA, human leukocyte antigen; NOS, not otherwise specified
*$p < 0.05$ compared to the other antibody subtypes

**Supplementary eTable6 Myopathology patterns and HLA-DR expression pattern 4 and 5**

| | Anti-ARS antibody | | | | | | | |
|---|---|---|---|---|---|---|---|---|
| | **All ARS** (n = 210)[a] | **Jo-1** (n = 65) | **OJ** (n = 20) | **PL-7** (n = 20) | **PL-12** (n = 11) | **EJ** (n = 10) | **KS** (n = 1) | **ARS_NOS** (n = 83)[a] |
| **Pattern 4** | | | | | | | | |
| Normal/non-specific | 6 (2.9) | 3(4.6) | 0 | 0 | 0 | 0 | 0 | 3 (3.6) |
| Necrotizing myopathy without PFN | 29 (13.8) | 14 (21.5) | 2 (10.0) | 1 (5.3) | 2 (18.2) | 0 | 0 | 10 (12.0) |
| Necrotizing myopathy with PFN | 24 (11.4) | 12 (18.5) | 2 (10.0) | 2 (10.5) | 0 | 2 (18.2) | 0 | 6 (7.2) |
| Others | 1 (0.5) | 1 (1.5)[b] | 0 | 0 | 0 | 0 | 0 | 0 |
| **Pattern 5** | | | | | | | | |
| Normal/non-specific | 1 (0.5) | 0 | 0 | 0 | 0 | 1 (9.1) | 0 | 0 |
| Necrotizing myopathy without PFN | 7 (3.3) | 2 (3.1) | 1 (5.0) | 0 | 0 | 0 | 0 | 4 (4.8) |
| Necrotizing myopathy with PFN | 11 (5.2) | 7 (10.8) | 1 (5.0) | 0 | 0 | 0 | 0 | 3 (3.6) |
| Others | 0 | 0 | 0 | 0 | 0 | 0 | 0 | 0 |

Categorial data is reported as number and (percentage). *$p < 0.05$ compared to the other antibody subtypes
Abbreviations: ARS, anti-tRNA synthetase; NOS, not otherwise specified; PFN, perifascicular necrosis
[a]Two cases of ARS, nos were excluded from the analysis due to artifacts
[b]Neurogenic muscle biopsy

**Supplementary eTable 7 HLA-DR expression patterns in non-ASS AIM and P-MM**

| | Non-ASS AIM (n = 602) | | | P-MM (n = 140) | | | | | |
|---|---|---|---|---|---|---|---|---|---|
| | **DM** (n = 188) | **IMNM** (n = 313) | **IBM** (n = 101) | **DYSF** (n = 50) | **SGP** (n = 15) | **LMNA** (n = 16) | **ANO5** (n = 3) | **FKRP** (n = 9) | **FSHD** (n = 47) |
| HLA-DR expression | 28 (14.9)* | 20 (6.4)* | 99 (98.0)* | 0 | 0 | 0 | 0 | 0 | 1 (2.1)* |
| Pattern 1 | 3 (1.6) | 11 (3.5) | 30 (29.7) | 0 | 0 | 0 | 0 | 0 | 1 (2.1) |
| Pattern 1+ | 0 | 0 | 65 (64.4) | 0 | 0 | 0 | 0 | 0 | 0 |
| Pattern 2 | 4 (2.1) | 3 (1.0) | 1 (1.0) | 0 | 0 | 0 | 0 | 0 | 0 |
| Pattern 3 | 7 (3.7) | 2 (0.6) | 0 | 0 | 0 | 0 | 0 | 0 | 0 |
| Pattern 4 | 10 (5.3) | 4 (1.3) | 0 | 0 | 0 | 0 | 0 | 0 | 0 |
| Pattern 5 | 4 (2.1) | 0 | 3 (3.0) | 0 | 0 | 0 | 0 | 0 | 0 |
| "Possible" perifascicular pattern (3+4+5) | 21 (11.2) | 6 (1.9) | 3 (3.0) | 0 | 0 | 0 | 0 | 0 | 0 |
| Perifascicular pattern (4+5) | 14 (7.4) | 4(1.3) | 3 (3.0) | 0 | 0 | 0 | 0 | 0 | 0 |

Categorial data is reported as number and (percentage).

**Abbreviations:** AIM, autoimmune myositis; ASS, antisynthetase syndrome; P-MM, possible myositis mimics; DM, dermatomyositis; IMNM, immune mediated necrotizing myopathy; IBM, inclusion body myositis; DYSF, dysferlinopathy; SGP, sarcoglycanopathy; LMNA, laminopathy; ANO5, anoctamin5 myopathy; FKRP, fukutin-related protein myopathy; FSHD, facioscapulohumeral disease

*$p < 0.05$ compared to ASS

**Supplementary eTable 8 Sensitivity, specificity, positive- and negative predictive value of HLA-DR expression in ASS**

| | **ASS (n=212)** | **Non-ASS[a] (n=742)** | **Sensitivity** | **Specificity** | **PPV** | **NPV** |
|---|---|---|---|---|---|---|
| HLA-DR expression | 128 (60.4) | 148 (19.9) | 60.4% | 80.1% | 46.4% | 87.6% |
| Considering ASS vs other entities excluding MxA-positive muscle biopsies and muscle biopsies from patient clinico-pathologically compatible with IBM | | | | | | |
| | **ASS[b] (n=209)** | **Non-ASS[c] (n=453)** | **Sensitivity** | **Specificity** | **PPV** | **NPV** |
| HLA-DR expression | 128 (61.2) | 21 (4.6) | 61.2% | 95.4% | 85.9% | 84.2% |
| HLA-DR 3+4+5 | 93 (44.5) | 6 (1.3) | 44.5% | 98.7% | 93.9% | 79.4% |
| HLA-DR 4+5 | 79 (37.8) | 4 (0.9) | 37.8% | 99.1% | 95.2% | 77.5% |
| Considering anti-Jo-1 ASS vs other entities excluding MxA-positive muscle biopsies and muscle biopsies from patient clinico-pathologically compatible with IBM | | | | | | |
| | **Jo-1 ASS (n=64)[d]** | **Non-Jo-1[c,e] (n=513)** | **Sensitivity** | **Specificity** | **PPV** | **NPV** |
| HLA-DR 3+4+5 | 41 (54.1) | 26 (5.1) | 64.1% | 94.9% | 61.2% | 95.5% |
| HLA-DR 4+5 | 39 (60.9) | 18 (3.5) | 60.9% | 96.5% | 68.4% | 95.2% |

[a]Non-ASS include DM=188, IMNM=313, IBM=101, P-MM=140
[b]Exclude 3 MxA-positive ASS: anti-Jo-1 = 1; anti-OJ =1; anti-PL-7=1
[c]Exclude DM=188 and IBM=101
[d]Exclude 1 MxA-positive anti-Jo-1 ASS
[e] Exclude 2 MxA-positive ASS: anti-OJ =1 and anti-PL-7=1
Abbreviation: HLA, human leukocyte antigen; ASS, antisynthetase syndrome; PPV, positive predictive value; NPV, negative predictive value; MxA, Myxovirus resistant protein A; DM, dermatomyositis; IMNM, immune mediated necrotizing myopathy; IBM, inclusion body myositis

**Supplementary eTable 9 Expression levels of the 10 most significantly expressed genes of the IFN-signaling pathway and *CIITA* in different subtypes of inflammatory myopathy**

| Jo-1 | | | OJ | | | IBM | | | HMGCR | | | SRP | | |
|---|---|---|---|---|---|---|---|---|---|---|---|---|---|---|
| **Gene** | **L2FC** | **padj** | **Gene** | **L2FC** | **padj** | **Gene** | **L2FC** | **padj** | **Gene** | **L2FC** | **padj** | **Gene** | **L2FC** | **padj** |
| *PSMB8* | 3.75 | 2.62E-51 | *PSMB8* | 3.22 | 1.89E-38 | *PSMB8* | 3.44 | 4.79E-36 | *IFI30* | 3.89 | 7.25E-16 | *IFI30* | 3.83 | 1.59E-15 |
| *B2M* | 2.76 | 1.04E-35 | *IFI30* | 5.45 | 1.32E-35 | *B2M* | 2.56 | 6.97E-26 | *FCGR1A* | 4.65 | 3.36E-11 | *TRIM14* | 2.59 | 8.26E-12 |
| *IRF9* | 2.68 | 2.04E-29 | *B2M* | 2.47 | 1.73E-29 | *HLA-A* | 2.63 | 2.16E-22 | *NCAM1* | 3.48 | 2.63E-10 | *PSMB8* | 1.76 | 2.38E-10 |
| *IRF1* | 3.51 | 7.17E-28 | *TRIM14* | 3.71 | 3.04E-27 | *GBP6* | 7.49 | 7.03E-22 | *IRF5* | 2.28 | 4.57E-10 | *IRF9* | 1.64 | 3.56E-10 |
| *HLA-A* | 2.61 | 2.39E-26 | *ICAM1* | 3.24 | 2.14E-25 | *HLA-F* | 2.88 | 4.48E-21 | *TRIM14* | 2.36 | 7.70E-10 | *IRF5* | 2.22 | 1.20E-09 |
| *ICAM1* | 3.35 | 2.94E-26 | *FCGR1A* | 6.54 | 5.99E-25 | *HLA-B* | 3.11 | 9.45E-21 | *OAS1* | 2.49 | 1.37E-09 | *OAS1* | 2.47 | 1.74E-09 |
| *GBP1* | 4.19 | 5.63E-26 | *IFI6* | 3.89 | 1.89E-21 | *HLA-E* | 1.84 | 5.18E-19 | *IRF8* | 2.79 | 4.69E-09 | *IRF7* | 1.96 | 3.22E-09 |
| *HLA-F* | 2.92 | 9.82E-26 | *SAMHD1* | 2.06 | 5.92E-21 | *HLA-DRA* | 3.68 | 1.55E-18 | *TRIM38* | 1.61 | 5.68E-09 | *IRF8* | 2.79 | 4.67E-09 |
| *IFI30* | 4.67 | 1.08E-25 | *HLA-A* | 2.25 | 1.75E-20 | *HLA-DPA1* | 3.38 | 1.64E-18 | *OASL* | 2.98 | 6.43E-09 | *OAS2* | 2.28 | 1.11E-08 |
| *HLA-B* | 2.87 | 4.27E-21 | *VCAM1* | 3.56 | 2.00E-20 | *HLA-DPB1* | 3.57 | 2.40E-18 | *HLA-DPA1* | 2.19 | 1.46E-08 | *VCAM1* | 2.42 | 1.91E-08 |
| *CIITA* | 2.43 | 6.53E-13 | *CIITA* | 1.80 | 5.71E-08 | *CIITA* | 2.58 | 2.10E-12 | *CIITA* | 1.29 | $7.40 \times 10^{-4}$ | *CIITA* | 1.04 | $6.75 \times 10^{-3}$ |

| TIF1-g | | | Mi-2 | | | MDA5 | | | NXP-2 | | |
|---|---|---|---|---|---|---|---|---|---|---|---|
| **Gene** | **L2FC** | **padj** | **Gene** | **L2FC** | **padj** | **Gene** | **L2FC** | **padj** | **Gene** | **L2FC** | **padj** |
| *ISG15* | 8.20 | 9.96E-63 | *IFI6* | 6.48 | 9.84E-54 | *ISG15* | 7.68 | 5.38E-66 | *ISG15* | 8.04 | 8.69E-93 |
| *IFI6* | 7.13 | 2.93E-61 | *OAS1* | 5.51 | 4.26E-48 | *IFI6* | 6.70 | 8.76E-65 | *IFI6* | 6.74 | 4.24E-84 |
| *IRF9* | 3.95 | 6.87E-57 | *IFITM1* | 3.68 | 2.38E-46 | *IRF9* | 3.85 | 8.76E-65 | *IFIT3* | 7.04 | 7.98E-81 |
| *IFIT3* | 7.15 | 3.16E-54 | *ISG15* | 6.81 | 2.47E-46 | *IFIT3* | 6.85 | 1.28E-59 | *OAS1* | 5.90 | 1.20E-79 |
| *OAS1* | 5.97 | 2.83E-53 | *MX2* | 4.60 | 1.08E-44 | *OAS1* | 5.55 | 5.88E-55 | *IFITM1* | 4.00 | 1.81E-79 |
| *RSAD2* | 5.93 | 1.45E-48 | *OAS3* | 4.89 | 2.36E-43 | *RSAD2* | 5.62 | 5.70E-52 | *IRF9* | 3.65 | 6.74E-75 |
| *IFITM1* | 3.78 | 3.35E-46 | *IFI27* | 4.27 | 2.08E-42 | *IFIT5* | 3.42 | 4.61E-50 | *OAS3* | 5.15 | 2.05E-69 |
| *OAS3* | 5.20 | 4.08E-46 | *OAS2* | 4.91 | 1.10E-40 | *OASL* | 6.57 | 9.90E-50 | *RSAD2* | 5.67 | 3.57E-68 |
| *IFIT5* | 3.55 | 3.55E-45 | *IFI30* | 5.93 | 2.77E-39 | *OAS3* | 4.85 | 5.79E-48 | *IFIT5* | 3.52 | 6.05E-68 |
| *PSMB8* | 3.71 | 7.04E-45 | *IFITM3* | 3.31 | 4.15E-39 | *IFITM1* | 3.51 | 1.65E-47 | *PSMB8* | 3.69 | 8.89E-68 |
| *CIITA* | 0.96 | $1.72 \times 10^{-2}$ | *CIITA* | 1.07 | $2.93 \times 10^{-3}$ | *CIITA* | 0.35 | 0.44 | *CIITA* | 1.02 | $7.67 \times 10^{-4}$ |

**Supplementary eTable 10 Pathology domains in antisynthetase syndrome of the original cohort**

| | Anti-ARS antibody | | | | | | |
|---|---|---|---|---|---|---|---|
| | **All ARS** (n = 50) | **Jo-1** (n = 15) | **OJ** (n = 13) | **PL-7** (n = 12) | **PL-12** (n = 4) | **EJ** (n = 5) | **KS** (n = 1) |
| **Muscle fiber domain** | | | | | | | |
| score | 3.3±2.2 | 2.2±1.3* | 5.2±2.0* | 3.3±2.5 | 1.3±0.5* | 3.6±2.4 | 4.0 |
| **Inflammatory domain** | | | | | | | |
| score | 4.5±2.9 | 3.9±2.9 | 7.2±3.0* | 3.9±1.8 | 2.8±1.0* | 2.8±2.4 | 2.0 |
| **Vascular domain** | | | | | | | |
| Capillary: muscle fiber ratio | 0.8±0.3[a] | 0.9±0.5 | 0.9±0.2[b] | 0.8±0.4[b] | 0.7 ±0.1 | 0.7 ±0.2 | 0.8 |
| **Connective tissue domain** | | | | | | | |
| PM-Fr | 27 (54.0) | 6 (40) | 9 (69.2) | 7 (58.3) | 3 (75.0) | 2 (40.0) | 0 |
| PM-ALP | 29 (58.0) | 6 (40) | 10 (76.9) | 8 (66.7) | 0* | 4 (80.0) | 1 (100.0) |
| Endomysial fibrosis | 3 (6.0) | 0 | 1 (7.7) | 1 (8.3) | 0 | 1 (20.0) | 0 |

Continuous data is shown as mean and ± standard deviation while categorial data is reported as number and (percentage).
Abbreviations: ARS, anti-tRNA synthetase; PM-Fr, perimysial connective tissue fragmentation; PM-ALP, increased perimysial alkaline phosphatase activity
[a]Two cases were excluded from analysis due to artifacts
[b]One case was excluded from analysis due to artifacts

**Supplementary eTable 11 Immunohistochemical feature in antisynthetase syndrome of the original cohort**

| | Anti-ARS antibody | | | | | | |
|---|---|---|---|---|---|---|---|
| | **All ARS** (n = 50) | **Jo-1** (n = 15) | **OJ** (n = 13) | **PL-7** (n = 12) | **PL-12** (n = 4) | **EJ** (n = 5) | **KS** (n = 1) |
| HLA-ABC expression | 49 (98.0) | 14 (93.3) | 13 (100.0) | 12 (100.0) | 4 (100.0) | 5 (100.0) | 1 (100.0) |
| HLA-ABC expression with PF enhancement | 2 (4.0) | 2 (13.3) | 0 | 0 | 0 | 0 | 0 |
| HLA-DR expression | 27 (54.0) | 9 (60.0) | 8 (61.5) | 4 (33.3) | 2 (50.0) | 3 (60.0) | 1 (100.0) |
| HLA-DR expression with perifascicular pattern (4+5) | 16 (32.0) | 8 (53.3)* | 4 (30.8) | 0* | 2 (50.0) | 2 (40.0) | 0 |
| MAC capillary deposition | 28 (56.0) | 12 (80.0)* | 7 (53.8) | 5 (41.7) | 2 (50.0) | 2 (40.0) | 0 |
| MAC capillary deposition in PF area | 2 (4.0) | 1(6.7)* | 1 (7.7)* | 0* | 0* | 0* | 0 |
| MAC sarcolemmal deposition | 19 (38.0) | 4 (26.7) | 7 (53.8) | 5 (41.7) | 0 | 3 (60) | 0 |
| MAC sarcolemmal deposition in PF area | 14 (28.0) | 2 (13.3) | 6 (46.2) | 4 (33.3) | 0 | 2 (40.0) | 0 |
| MxA expression | 1 (2.0) | 0 | 0 | 1 (8.3) | 0 | 0 | 0 |

Continuous data is shown as mean and ± standard deviation while categorial data is reported as number and (percentage).
Abbreviations: ARS, anti-tRNA synthetase; HLA, human leukocyte antigen; PF, perifascicular area; MAC, membrane attack complex; MxA, myxovirus resistant protein A
*p < 0.05 compared to the other antibody subtype

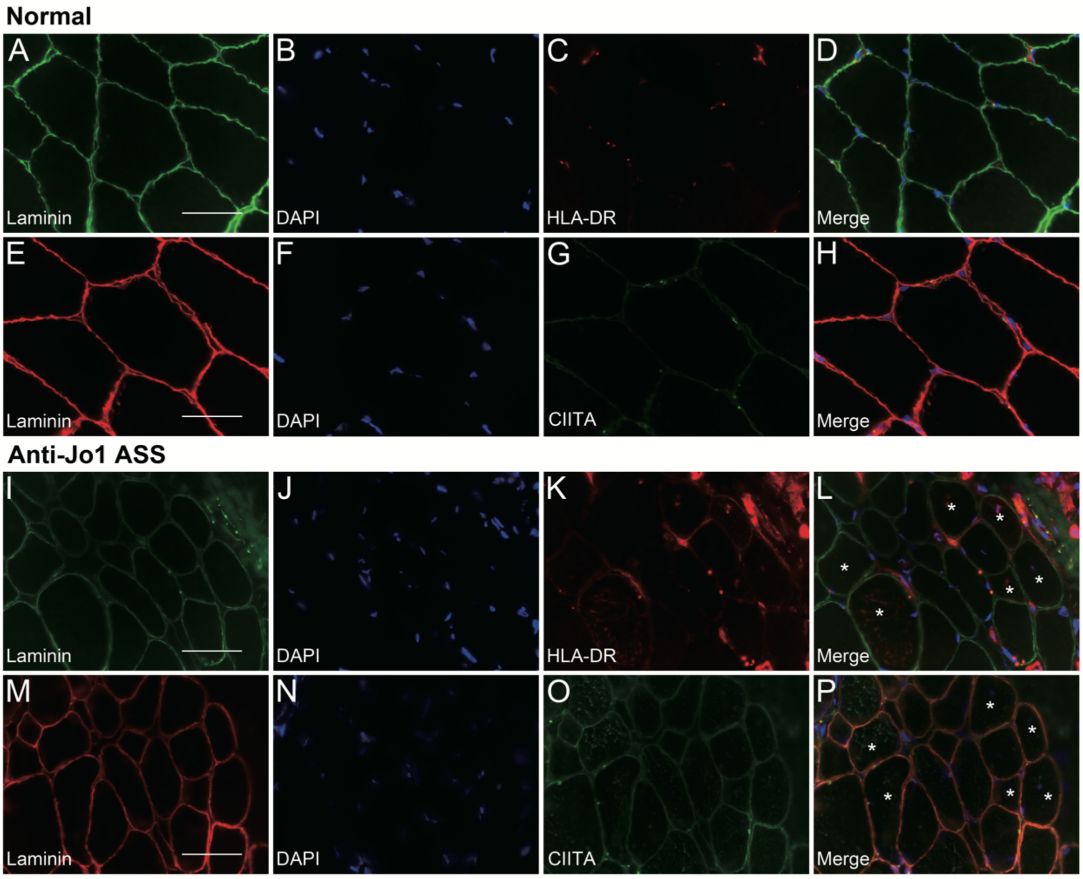


**Supplementary efigure 1 CIITA and HLA-DR expression**

Normal muscle biopsy **(A-H):** Staining for HLA-DR and CIITA are negative in normal muscle biopsy. Serialized section of anti-Jo-1 ASS **(I-P):** Some fibers show HLA-DR and CIITA expression; co-expression is focally observed (white asterisk, **L and P**).

**Note:** A-P bar = 50μm; A, E, I, M: laminin; B, F, J, N: DAPI; C, K: HLA-DR (human leukocyte antigen-DR, major histocompatibility complex class II); G, O: CIITA (major histocompatibility complex class II activator); D,H,L,P: merge.
